# The causal relationship between Graves’ disease and type 2 diabetes: Evidence from Bidirectional Mendelian Randomization Study

**DOI:** 10.1101/2023.11.07.23296920

**Authors:** Mahdi Akbarzadeh, Mahsa Heidari-Foroozan, Samaneh Talebi, Danial Habibi, Sahand Tehrani Fateh, Farideh Neshati, Majid Valizadeh, Amir Hossein Ghanooni, Hamid Alavi Majd, Parisa Riahi, Fereidoun Azizi, Mehdi Hedayati, Maryam Sadat Daneshpour

**Affiliations:** Cellular and Molecular Endocrine Research Center, Research Institute for Endocrine Sciences, Shahid Beheshti University of Medical Sciences, Tehran, Iran.; Student Research Committee, School of Medicine, Shahid Beheshti University of Medical Sciences, Tehran, Iran.; Department of Biostatistics, School of Allied Medical Sciences, Shahid Beheshti University of Medical Sciences, Tehran, Iran.; Department of Biostatistics and Epidemiology, School of Health, and Student Research Committee, School of Health, Isfahan University of Medical Sciences, Isfahan, Iran.; School of Medicine, Tehran University of Medical Sciences, Tehran, Iran.; Obesity Research Center, Research Institute for Endocrine Sciences, Shahid Beheshti University of Medical Sciences, Tehran, Iran.; Department of Endocrinology, School of Medicine, Iran University of Medical Sciences, Tehran, Iran.; Endocrine Research Center, Research Institute for Endocrine Sciences, Shahid Beheshti University of Medical Sciences, Tehran, Iran.

**Author notes:** **Corresponding author: Maryam Sadat Daneshpour** (Ph.D.), Associate Professor. Cellular and Molecular Endocrine Research Center, Research Institute for Endocrine Sciences, Shahid Beheshti University of Medical Sciences.; P.O. Box: 19395-4763, 1985717413.

**Keywords:** Diabetes Mellitus, Graves’ Disease, Mendelian Randomization Analysis, T2D

## Abstract

**Introduction:** Numerous observational investigations have hinted at a possible link between Graves’ disease (GD) and the susceptibility to Type 2 diabetes (T2D). The primary objective of this study was to explore the potential underlying causal connection between GD and T2D through the application of bidirectional Mendelian randomization (MR) analysis.

**Materials and Methods:** MR analysis was conducted with summary-level data from genome-wide association studies (GWAS) for GD and T2D. Single-nucleotide polymorphisms (SNPs) for GD were extracted from 458,620 (1,678 cases and 456,942 controls) Europeans in the NBC Human Database, and the summary-level data of T2D (180,834 cases and 1,159,055 controls) were extracted from the DIAGRAM consortium.

The primary analysis method was the inverse variance weighted (IVW) method. Heterogeneity and pleiotropy were considered to assess the causal relationship between GD and T2D. Sensitivity analyses were conducted to ensure the robustness of the findings.

**Results:** The initial analysis found no significant causal relationship between GD and an increased risk of T2D (OR = 1.019, 95% CI (0.997,1.042); SE = 0.01, P= 0.373). However, after sensitivity analyses and removal of outlier SNPs, a significant causal relationship was found (OR_IVW_ = 1.017, 95% CI (1.002,1.033), P= 0.03, Cochran’s Q= 7.8, p-value = 0.932, I^2^ = 0.0%).

**Conclusions:** The study indicates a causal link between GD and an elevated T2D risk, underscoring the need for blood sugar monitoring and specialized care for GD patients. Further research into GD-T2D mechanisms is essential for preventive strategies and interventions.

## Introduction

Diabetes mellitus (DM) stands as a prevalent global health challenge, affecting approximately 9% of the global populace, with nearly 90% of cases attributed to type 2 diabetes (T2D). Furthermore, an alarming 22.9 million new T2D cases surfaced worldwide in 2017 (1). Controlling the increasing T2D burden requires an insightful comprehension of its underlying etiology and risk factors (1). Thyroid hormones assume a pivotal role in metabolic regulation, exerting both direct and indirect influences on insulin dynamics including production, secretion, and clearance. Notably, hyperthyroidism engenders hyperglycemic states (2). Graves’ disease (GD), an autoimmune disorder characterized by antibodies targeting the thyrotropin receptor, is the leading cause of hyperthyroidism. Observational studies have reported the comorbidity of thyroid dysfunction and diabetes mellitus, with prevalence rates of thyroid dysfunction ranging from 10.8% to 13.4% within DM cohorts (3). Interestingly, a notable subset of hyperthyroidism cases transitions into DM. Nevertheless, the causal relationship between GD and DM remains yet to be elucidated, with most of the studies reporting the prevalence of thyroid dysfunction in DM cohorts (2–4).

Observational studies, as is inherent to their design, are susceptible to various biases, including confounding factors, study design intricacies, and the complexities of reverse causality (5). Considering the scarcity of conclusive causal evidence linking GD and T2D, the innovative approach of Mendelian randomization (MR) holds the potential to furnish insight. MR efficiently utilizes the genetic diversity that naturally affects thyroid function and the likelihood of developing T2D. This basis is reinforced by genetic elements that account for approximately 65% of the detected variations in Thyroid-Stimulating Hormone (TSH) and Free Thyroxine (Free T4) levels (6).

This study uses the results of Genome-Wide Association Study (GWAS) to investigate the causal interplay between GD and T2D. In parallel, we also investigate the reciprocal relationship, exploring whether T2D increases vulnerability to GD. To the best of our knowledge, this is the first study to investigate the causal relationship between GD and T2D using the MR approach.

## Materials and Methods

### Study design

We were used a bidirectional MR analysis. Due to Figure 1, it is essential to assess the validity of these assumptions in every MR analysis(7): 1) The SNPs chosen as instrumental variables (IVs) should have a strong association with exposure. 2) The IVs should not be related to the confounding factors that influence outcome and exposure association. 3) The effect of the IVs on outcome should be solely through their impact on exposure and not through a direct association with outcome.

**Figure 1:**
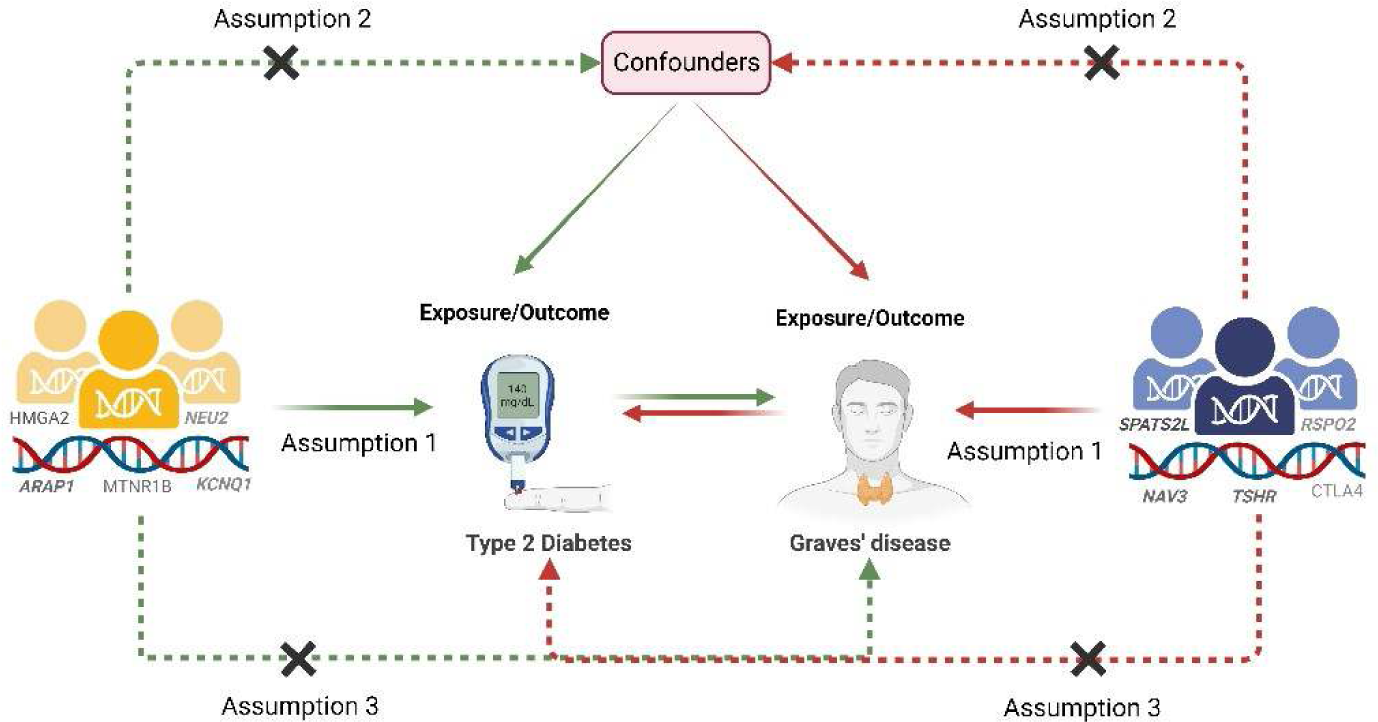
Mendelian randomization is a statistical technique used in epidemiology and genetics to assess causal relationships between an exposure (e.g., a risk factor) and an outcome (e.g., a disease). It relies on genetic variants that are randomly inherited at conception to mimic the randomization process of a controlled experiment, helping researchers infer causal relationships. Figure 1 shows the framework of two sample bidirectional Mendelian randomization analysis is based on several key assumptions. 1: The genetic variations are strongly associated with exposure; 2: The genetic variations are not associated with either known or unknown confounders; 3: SNPs should influence risk of the outcome through the exposure, not through other pathways. The green line represented the Mendelian randomization analysis of the association of GD with T2D. The red line represented the Mendelian randomization analysis of the association of T2D with GD.

MR analysis was employed to investigate the causal relationship between GD and T2D. GD as an autoimmune disorder that leads to overactivity of the thyroid gland (hyperthyroidism), and T2D as a type of chronic metabolic disorder of insulin resistance or desensitization and increased blood glucose levels were assessed in this study.

The summary-level data for bidirectional MR analysis were obtained from the GWAS catalog database, namely Graves’ disease (GCST90018847) and Type 2 diabetes (GCST90132184). The details of summary-level data for each GWAS are available in Table 1.

**Table1.** Detailed information of GWAS summary statistics data used for analyses.

|  | Phenotype | Reporting traits in the database | Source | GWAS ID | Sample size | Ethnicity | Reference |
| --- | --- | --- | --- | --- | --- | --- | --- |
| **exposure** | Graves' disease | Graves' disease | GWAS catalog | (GCST90018847) | 1,678 (case),<br>456,942 (control) | European | Sakaue et al. 2021 |
| **outcome** | Type 2 diabetes | Type 2 diabetes | GWAS catalog | (GCST90132184) | 180,834 (case),<br>1,159,055 (control) | European (51%),<br>East Asian (28%),<br>South Asian (8%),<br>African (7%),<br>Hispanic (6%) | Mahajan et al. 2022 |

### Instrumental SNPs selection

For extracting significant SNPs for GD, we used a relaxed GWAS threshold of p − value < 5 × 10^-5^(8, 9). On the other hand, for extracting significant SNPs associated with T2D we employed a more stringent genome-wide threshold of p − value < 5 × 10^-8^(10).

In the final sets, we performed linkage disequilibrium (LD) clumping restricted to r^2^ < 0.001 in a clumping distance = 10,000 kb window to minimize correlations between the selected SNPs. After clumping, SNPs that were missed in the outcome GWAS dataset were replaced with SNPs from the NIH LDproxy database according to EUR ancestry (11, 12). The exposure and outcomes data were harmonized to ensure alleles were aligned, and the presence of ambiguous and palindromic variants was checked. Additionally, SNPs with a minor allele frequency (MAF) less than 0.01 were omitted. We checked PhenoScanner results for the SNPs used to detect potential confounders (13, 14).

The strength of each instrumental variable (IV) was evaluated using F-statistics, which is calculated as 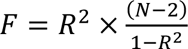, *where* 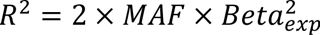, represents the total variance of the extracted SNPs, and N is the sample size. IVs with statistical value (F > 10) were included (15). Detailed information on the SNPs used is presented in Supplementary Table S1 and S2.

### Statistical analysis

The primary analysis method used was inverse-variance weighted (IVW) method. The effect size was reported as an odds ratio (OR) with a 95% confidence interval (CI). In addition, several other methods were employed in this study to examine the stability and reliability of the results, including Penalized MR-Egger, Robust MR-Egger, Penalized robust MR-Egger, Simple median, Penalized weighted median, Simple Mode, Weighted Mode, Penalized IVW, Robust IVW, Penalized robust IVW, MR-Lasso, MR-Constrained maximum likelihood (MR-cML), Debiased inverse-variance weighted, and Model-based estimation (MBE) method (16).

Heterogeneity in the analysis was addressed by using multiple methods, including Cochran’s Q statistic, the I^2^ index, and Rucker’s Q statistic (17). Through funnel plots outlier SNPs were identified. Horizontal pleiotropy was assessed through intercept tests using the MR-Egger method (18). To find and eliminate pleiotropic SNPs, the human gene phenotypic association database, Phenoscanner (http://www.phenoscanner.medschl.cam.ac.uk), was scanned. The MR pleiotropy Residual Sum and Outlier (MR-PRESSO) test was conducted to detect possible outliers and correct for horizontal pleiotropy by removing outliers (19).

Cook’s distance was used to diagnose and eliminate SNPs that had a significant and false impact on the model’s fitted values (18). SNPs identified as outliers and influential points were excluded from further MR analysis. Sensitivity analyses, such as leave-one-out and single SNP MR, were also performed to assess the robustness of the results (17).

All analyses were performed using R packages “*TwoSampleMR*”, “*MendelianRandomization*”, “*MRPRESSO*”, and “*mr.raps*” in R software 4.3.1 implemented in R Studio (20, 21).

## Results

### SNPs selection

As shown in Figure 2A, 11,797 SNPs passed the genome-wide significance threshold (p − value < 5 × 10^-5^) in GD GWAS. Six of the 31 SNPs remaining after clumping were missing in the T2D GWAS dataset. 3 SNPs (rs793102, rs1049053, and rs144334834 with R^2^ 0.99, 0.91, and 0.94, respectively) were chosen from the NIH LDproxy database based on EUR ancestry instead of those SNPs. The process of MR analyses and the results are publicly available through the following HTML link: https://akbarzadehms.github.io/GDT2D-MR/

**Figure 2:**
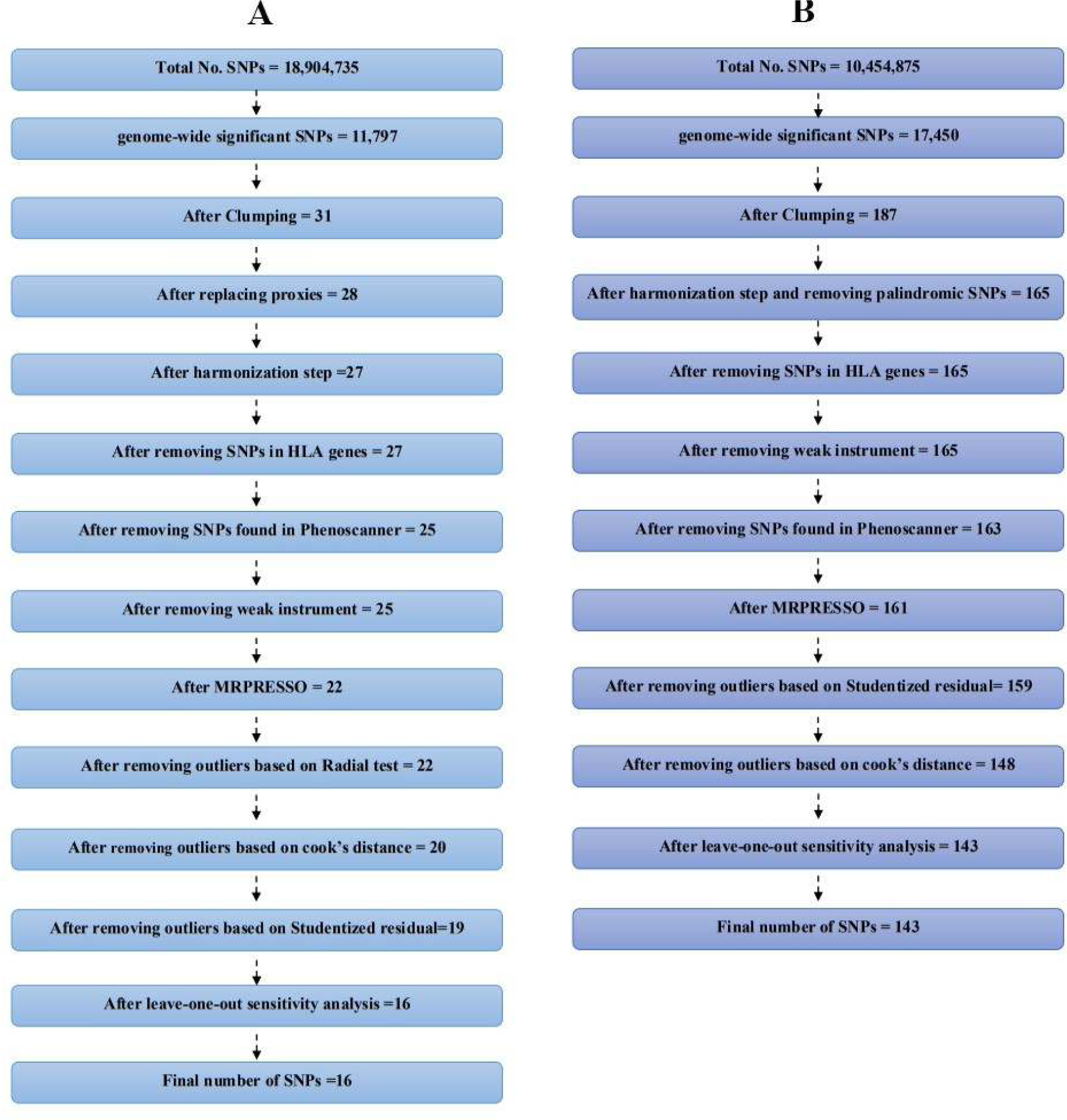
Selecting an appropriate instrumental variable (IV) in Mendelian randomization is a crucial step because the validity of your causal inference depends on it. Here are the key steps involved in the selection of an IV in Mendelian randomization. Genome-Wide Significant SNP: In this step, researchers identify a genetic variant (SNP) that is genome-wide significant in genome-wide association studies (GWAS) for the exposure of interest. Genome-wide significance means that the SNP is highly associated with exposure. Clumping: Clumping involves selecting a subset of SNPs in linkage disequilibrium with the primary SNP. Replacing Proxies: Sometimes, the initial genome-wide significant SNP is not qualified as an instrumental variable due to issues like pleiotropy or linkage disequilibrium with other SNPs. In such cases, the primary SNP is replaced with a proxy SNP that has a similar effect on the exposure but fewer issues. Harmonization Step: Data harmonization is the process of standardizing genetic data from different sources or studies to ensure compatibility. This step is crucial when combining data from multiple datasets. Removing HLA Gene: The HLA (Human Leukocyte Antigen) gene region is known for its extensive genetic diversity and complex associations with various traits. In MR analysis, it’s common to exclude SNPs in the HLA region to minimize potential confounding or pleiotropic effects. Check Phenoscanner: Phenoscanner is a tool used to explore genetic associations with various phenotypes, which can provide insights into pleiotropy. Removing Weak Instrument: It’s crucial to assess the strength of the selected genetic instrument(s) by checking for the F-statistic or MAF value. Weak instruments can lead to imprecise estimates, so we removed them from the analysis. Detect outliers: MRPRESSO and the Radial Test aim to improve the robustness of Mendelian randomization by identifying and addressing potential sources of bias due to pleiotropy. By detecting and correcting for outliers, these methods help ensure more accurate causal inference in MR analyses. Additionally, SNPs with high Cook’s distances were checked more closely. If these SNPs are identified as influential outliers, they are candidates for exclusion from the MR analysis. Finally, it’s essential to use a combination of statistical techniques and biological knowledge to make informed decisions about which SNPs to include or exclude in MR analyses.

The harmonization process resulted in the exclusion of one SNP due to incompatible alleles. After conducting pleiotropy checks using PhenoScanner, 25 SNPs remained for further analysis. The individual F-statistics ranged from 19.62 to 109.46 (Supplementary Table S1).

### The causal effect of GD on the risk of T2D

With 25 SNPs as IVs, the results of IVW showed no significant causal relationship between GD a nd T2D (OR = 1.019, 95% CI (0.997,1.042); SE = 0.01, P= 0.373). According to the pleiotropy test, there was no pleiotropy (MR–Egger intercept p value = 0.096). But heterogeneity was found (t he Q-p values of IVW and MR–Egger were 1.895×10^-6^ and 1.798×10^-5^, respectively).

### Sensitivity analysis

In sensitivity analysis, three, three, four, one, and five SNPs were detected as outliers or influential observations using MR-PRESSO, Radial test, leave-one-out sensitivity analysis, Studentized residual, and Cook’s distance, respectively. Considering that these methods overlap each other, fina lly 9 SNPs were excluded (highlighted in Supplementary Table S1).

After excluding those SNPs, 16 remained IVs for further MR analysis. Neither pleiotropy (MR–Egger intercept p-value = 0.819) nor heterogeneity (Cochran’s Q= 7.8, p-value = 0.932, /^2^ = 0.0%) were detected in the analysis. Therefore, based on the results of IVW (OR_IVW_ = 1.017, 95% CI (1.002,1.033), P= 0.03), Penalized IVW (OR _Penalized IVW_ = 1.017, 95% CI (1.002,1.033), P= 0.03), MR-RAPS (OR _MR-RAPS_ = 1.017, 95% CI (1.001,1.034), P= 0.036), MR-Lasso (OR _MR-Lasso_ = 1.01 7, 95% CI (1.002,1.033), P= 0.03), MR-cML (OR _MR-cML_ = 1.017, 95% CI (1.002,1.034), P= 0.03 1), and Debiased inverse-variance weighted (OR _dIVW_ = 1.018, 95% CI (1.002,1.035), P= 0.031), GD was recognized as a risk factor for T2D. In other MR methods, there was no significant causal relationship between GD and T2D (the p-values of models exceeded 0.05).

The forest plot in Figure 3A and scatter plots in Figure 4A present the results of other MR methods. Supplementary Figures S1B, S1C, and S1D provide funnel plot, leave-one-out sensitivity analysis, and the single SNP sensitivity analysis, respectively.

**Figure 3:**
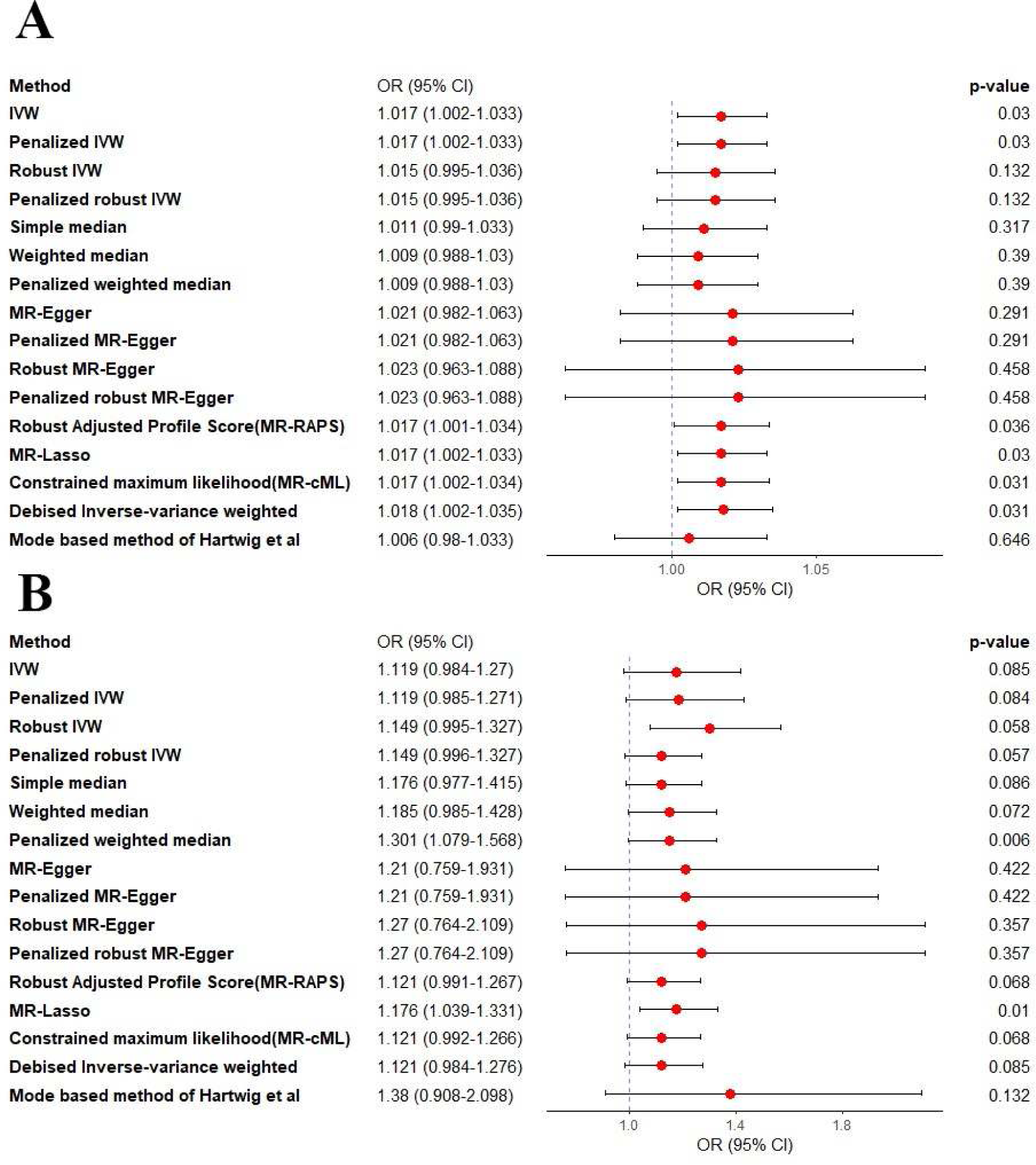
A forest plot of multiple methods in Mendelian randomization (MR) is a graphical representation that displays the results of different MR approaches used to estimate causal effects. On the left side of the forest plot, we listed the various MR approaches we have applied. According to forest plot A, based on the results of IVW (ORIVW = 1.017, 95% CI (1.002,1.033), P= 0.03), Penalized IVW (OR Penalized IVW = 1.017, 95% CI (1.002,1.033), P = 0.03), MR-RAPS (OR MR-RAPS = 1.017, 95% CI (1.001,1.034), P = 0.036), MR-Lasso (OR MR-Lasso = 1.017, 95% CI (1.002,1.033), P = 0.03), MR-cML (OR MR-cML = 1.017, 95% CI (1.002,1.034), P= 0.031), and Debiased inverse-variance weighted (OR dIVW = 1.018, 95% CI (1.002,1.035), P= 0.031), GD was recognized as a risk factor for T2D. According to forest plot B, results of IVW (ORIVW = 1.118, 95% CI (0.987,1.271), P= 0.085), Penalized IVW (OR Penalized IVW = 1.119, 95% CI (0.985,1.271), P= 0.304), Robust IVW (OR Robust IVW = 1.149, 95% CI (0.995,1.327), P= 0.182) and Penalized Robust IVW (OR Penalized Robust IVW = 1.149, 95% CI (0.996,1.327), P= 0.183), T2D had no significant causal effect on the risk of GD.

**Figure 4:**
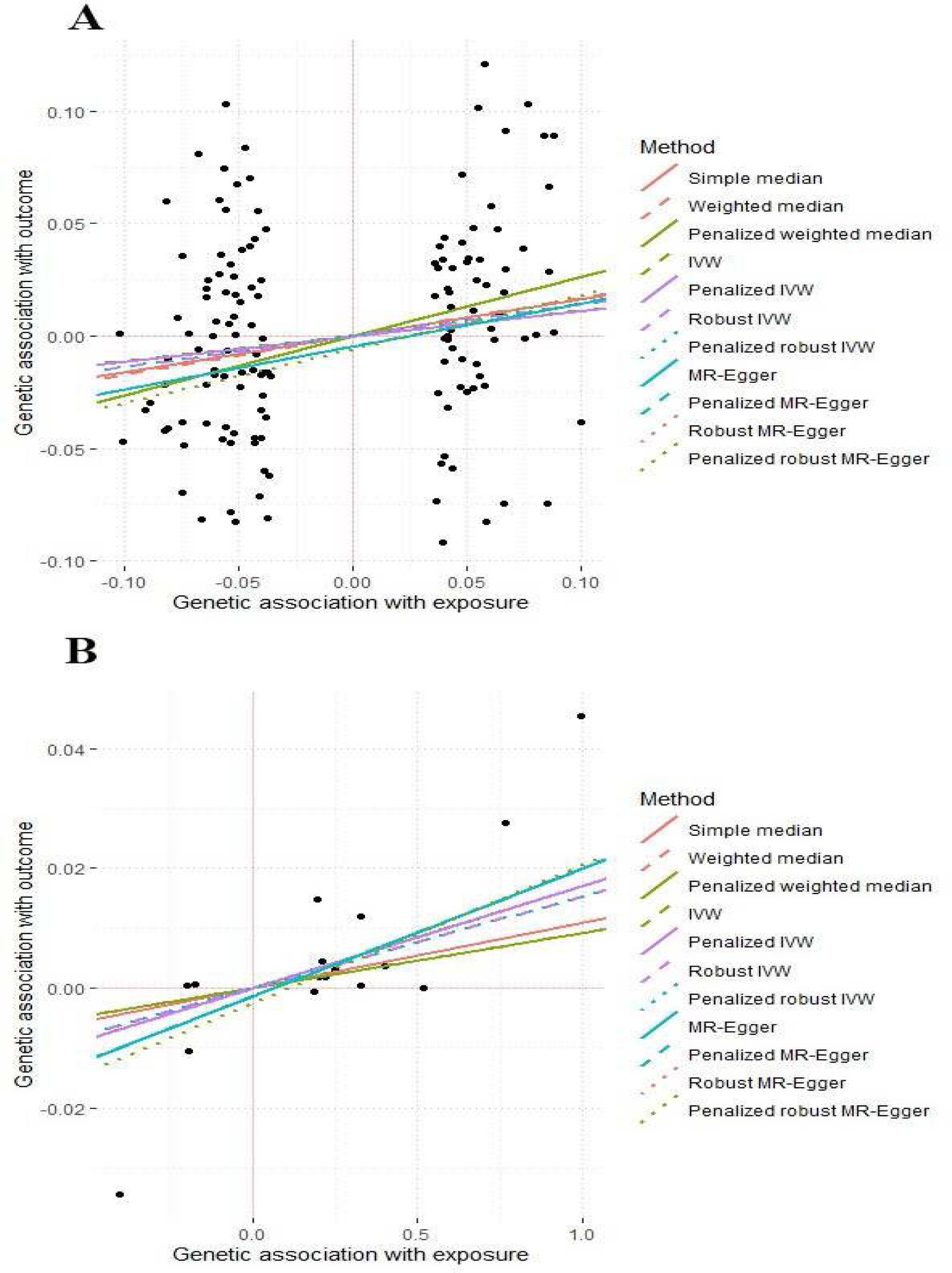
A scatter plot of various methods in Mendelian randomization (MR) is a graphical representation used to visualize and compare the results of different MR approaches. This type of plot allows us to examine the relationships and agreements (or disagreements) in the causal effect estimates obtained by applying multiple MR methods. Scatter plots A of the effect of GD-association SNPs on T2D exhibit positive causality about GD on T2D.

### The causal effect of T2D on the risk of GD

To assess the causal effect of T2D on the risk of GD, as shown in Figure 2B, 17,450 SNPs passed the genome-wide significance threshold (p − value < 5 × 10^-8^) in T2D GWAS. 17 of the 187 SNPs remaining after clumping were missing in the GD GWAS dataset.

5 SNPs of 170 were removed for being palindromic in the harmonization step. To ensure the absence of pleiotropy, we used PhenoScanner to assess pleiotropic effects, resulting in the selection of 163 SNPs as genetic instruments. Individual F-statistics ranged from 29.75 to 1415.77 (Supplementary Table S2).

With 163 SNPs as IVs, the results of IVW showed significant causal relationship between T2D a nd GD (OR = 1.14, 95% CI (1.012,1.274), P= 0.031). According to the pleiotropy test, there was no pleiotropy (MR–Egger intercept p value = 0.974). But heterogeneity was found (the Q-p value s of IVW and MR–Egger were 1.41×10^-3^ and 1.19×10^-3^, respectively).

### Sensitivity analysis

The results of sensitivity analysis led to the removal of 20 SNPs, which are highlighted in Supplementary Table S2. Consequently, 143 IVs were utilized for MR analysis. According to the results of IVW (OR_IVW_ = 1.118, 95% CI (0.987,1.271), P= 0.085), Penalized IVW (OR _Penalized IVW_ = 1.119, 95% CI (0.985,1.271), P= 0.304), Robust IVW (OR _Robust IVW_ = 1.149, 95% CI (0.995,1.327), P= 0.182) and Penalized Robust IVW (OR _Penalized Robust IVW_ = 1.149, 95% CI (0.996,1.327), P= 0.183), T2D had no significant causal effect on the risk of GD. No evidence of pleiotropy was found, as indicated by the MR-Egger intercept (p-value =0.73). Additionally, no heterogeneity (Cochran’s Q=158.491, p-value =0.163, I^2^ = 10.4%) was observed in the analysis. Furthermore, the p-values obtained from the weighted median, weighted mode, and MR-Egger methods exceeded 0.05. The forest plot in Figure 3B and scatter plots in Figure 4B present the results of other MR methods, while supplementary Figures S2B, S2C, and S2D provide funnel plot, leave-one-out sensitivity analysis, and the single SNP sensitivity analysis, respectively.

## Discussion

Comorbidity is a common situation in clinical practice where certain medical conditions are frequently observed together. This often leads to the simultaneous identification of these co-occurring diseases by healthcare professionals. When diseases are comorbid, it suggests the possibility that they may have common causes and mechanisms. This understanding is particularly significant for determining appropriate treatment approaches for these interconnected diseases. In this study, we examined the potential cause-and-effect relationship between GD and T2D using the MR technique. The results indicated that GD, as a distinct medical condition, plays a causal role in the development of T2D. However, the bilateral relationship does not exist and we could not find a causal effect between T2D on GD.

Genetic factors play a crucial role in the onset of various diseases. Previous research has demonstrated that the concordance rates, which indicate the likelihood of both monozygotic twins developing T2D can be remarkably high, reaching up to 76% (22). Moreover, extensive studies on GD involving twins have shown a significant concordance rate of 35% in monozygotic twins, in contrast to a mere 3% in dizygotic twins.

GWAS studies have shown a significant genetic susceptibility that overlaps across various tissues, particularly in the context of immune-related diseases like GD. Human leukocyte antigen (HLA) genes exert control over immune response-related genes, and the presence of HLA associations signifies their implication in autoimmunity. The HLA-DPA1*01:03-*01:03 and HLA-DPA1*01:03-02:01 alleles are associated with both T2D and GD (23). Additionally, HLA DRB103 is proposed as a shared causative factor in both GD and T2D (24). Furthermore, besides HLA genes, non-HLA genes also play an important role in development of these diseases. For instance, microsatellite polymorphisms in the PSMA6 is also associated with T2D on GD in previous research (25).

The acknowledgment of the connection between hyperthyroidism and DM can be traced back to 1927, when it was initially observed by Coller and Huggins. In their study, they documented a series of 12 cases with DM and hyperthyroidism. They noted that hyperthyroidism exacerbated hyperglycemia in these individuals, and significant improvement in managing blood sugar levels was observed following thyroidectomy (26).

The impact of thyroid hormones on glucose metabolism is well-established. Thyroid hormones play a direct and indirect role in regulating insulin secretion, glucose disposal and hepatic glucose output. In hyperthyroidism, many aspects of glucose metabolism are altered. Namely, the cleavage of C-peptide to pre-insulin is impaired (27) and the first response to glucose is lowered (28). Moreover, consensus exists concerning the impact of thyroid hormones on hepatic insulin resistance. Early experiments conducted with hyperthyroid rat models revealed their incapacity to synthesize glycogen and inhibit hepatic glucose production in the fed state (29). When compared to individuals with normal thyroid function (euthyroid patients), those with GD exhibit a significant 90% increase in fasting glucose production and a notable 50% reduction in the ability to suppress hepatic glucose production when insulin is administered (30).

It’s important to note that the binding of insulin to hepatic receptors remains unaltered in the context of thyrotoxicosis. This suggests that the hepatic insulin resistance observed in thyrotoxicosis occurs downstream of the insulin receptor (31). Furthermore, triiodothyronine (T3) directly stimulates hepatic lipogenesis, which appears to be independent of insulin’s influence. This stimulation contributes to a cycle of continuous lipogenesis and lipid oxidation, ultimately promoting the development of ketoacidosis (32).

However, it is noteworthy that in hepatocyte culture models exposed to excessive T3 levels, several insulin-dependent processes show decreased function. These include insulin-stimulated glycogen synthesis, insulin’s ability to suppress gluconeogenesis, and insulin-induced lipogenesis, independent of T3 effects, all of which are compromised, indicating a state of insulin resistance (33).

In adipose tissue, thyroid hormones (TH) have several effects. They enhance the process of lipid oxidation (34) and amplify the responsiveness of adipocytes to catecholamines, which in turn indirectly boosts the release of fatty acids (35).

Conversely, in skeletal muscles, TH has a different impact. It reduces the rate of glycogen synthesis and diverts glucose toward conversion into lactate, a process that contributes to the Cori cycle in the liver. Consequently, this redirection of glucose metabolism leads to an elevated output of glucose (36)

Despite the established evidence of insulin resistance in GD patients, the precise risk of developing new-onset DM in individuals with hyperthyroidism in real-world clinical practice remains uncertain There are only a few long-term studies examining the risk of DM in individuals with thyroid dysfunction with most of them primarily associating this risk with hypothyroidism rather than exploring its link to hyperthyroidism (37–39).

In our study, we found a causal effect between GD on T2D, our results were in line with that of Song *et al.* which showed a hazards ratio of 1.18 (95% confidence interval [CI], 1.10 to 1.28) for developing DM in individual with long-term GD (3). Moreover, Brandt *et al.* also reported an increased risk for DM in hyperthyroid individuals with a hazard’s ratio of 1.46 (95% CI, 1.16 to 1.84) (37).

Remarkably, current guidelines for managing hyperthyroidism do not include recommendations for monitoring blood sugar levels or screening for diabetes in hyperthyroid patients Considering that individuals with longstanding GD have a notably higher susceptibility to diabetes compared to the general population, it is crucial to provide specific care and regular monitoring for the emergence of DM in these patients, especially those who have experienced prolonged periods of hyperthyroidism (40, 41).

Although the mechanisms of which the GD may impact T2D has been documented in the literature, studies regarding the effect of T2D on GD is scarce. Diabetes may impair thyroid function by reducing TSH level and inhibiting the conversion of T4 to T3 in the peripheral tissues. Diabetic ketoacidosis could also lower the T4 and T3 level (4), and it worth mentioning that insulin resistance and hyperinsulinemia enhance thyroid issue proliferation and hence can cause goiter and also increase the incidence of nodular thyroid disease (42). Despite the effect of diabetes on thyroid function, the exact effect of T2D on GD is not well understood. The result of the present study also could not found any significant causal effect between T2D on GD (OR_IVW_ = 1.118, 95% CI (0.987,1.271), P= 0.085).

The identification of a causal effect between GD on T2D calls for attention for a comprehensive approach to clinical management. To address this, it is beneficial to implement regular blood sugar monitoring in individuals with GD as a standard practice. Simultaneously, healthcare professionals should emphasize lifestyle modifications, including dietary improvements and increased physical activity, which can help regulate blood sugar levels and reduce the risk of T2D development. Patient education about the GD-T2D connection is vital to promote proactive self-care. Long-term surveillance programs, can detect potential T2D cases early, enabling timely intervention. Further research on the mechanisms linking GD and T2D is crucial for developing targeted therapies and interventions that minimize the T2D risk in GD patients. These measures can not only lower T2D incidence but also enhance the overall quality of life of the individuals living with GD.

This study boasts several notable strengths. Firstly, it leverages data from well-established and extensively-utilized GWAS databases, which are renowned for their reliability and encompass considerable sample sizes. The use of such robust data sources enhances the study’s scientific rigor and the confidence in its findings. Secondly, the application of the two-sample MR method, where genetic variants serve as instrumental variables, contributes to the study’s methodological robustness. This approach effectively diminishes the influence of confounding factors, ensuring that the observed relationships are more likely to be causal in nature. Moreover, the study incorporates sensitivity analyses and evaluates pleiotropy, which are critical steps in strengthening the reliability of the results. By systematically assessing the possibility of bias from outliers and considering the multifaceted effects of instrumental variables, the study demonstrates a comprehensive and thorough research approach.

Despite these strengths, it is important to acknowledge certain limitations within the study. One notable limitation is the reliance on GWAS data primarily derived from European populations. While these data sources provide valuable insights, their generalizability to other ethnic groups may be restricted. As such, caution is warranted when applying the study’s findings to populations with different genetic backgrounds and environmental contexts. Additionally, the use of GWAS data introduces potential challenges related to population stratification and relatedness. Even though extensive efforts were made to account for these issues, there remains the possibility of residual bias stemming from these sources. It is crucial to recognize that despite the study’s robust methodology; the results should be interpreted with caution due to these limitations. Furthermore, due to the cross-sectional origin of this study, it cannot determine the effect of the duration of the clinical or subclinical hyperthyroidism on T2D and how the amount of time the disease stays in remission affects the causality of GD on T2D.

## Conclusion

In conclusion, we conducted a comprehensive bidirectional Mendelian randomization analysis to investigate the intricate relationship between GD and T2D. we reported a causal association, suggesting that GD increases the risk of developing T2D. These results highlight the clinical significance of closely monitoring blood sugar levels in individuals with GD and tailoring care accordingly.

Moreover, our research highlights the imperative for further investigations into the underlying mechanisms that link GD and T2D. A deeper understanding of these mechanisms will be pivotal in the development of targeted preventive strategies and interventions. By addressing this important health connection, we aim to enhance the quality of life and overall health outcomes for individuals living with GD.

## Data Availability

The original data used are publicly available at https://www.ebi.ac.uk/gwas/.

## Acknowledgments

We want to acknowledge the participants and investigators who made summary data available.

## Data Availability Statement

The original data used are publicly available at https://www.ebi.ac.uk/gwas/. The original contributions presented in the study are included in the article/Supplementary Material; The process of MR analyses and the results are publicly available through the following HTML link: https://akbarzadehms.github.io/GDT2D-MR/

## Disclosure

### Conflict of interest

All authors declare that they have no conflicts of interest.

Approval date of Registry and the Registration No. of the study/trial: The ethical committee approved this study at the Research Institute for Endocrine Sciences, Shahid Beheshti University of Medical Sciences (Research Approval Code: 28778 & Research Ethical Code: IR.SBMU.ENDOCRINE.REC.1400.084). In this study, all participants provided written informed consent for participating in the study. This study has been performed following the Declaration of Helsinki.

Approval of the research protocol: It is accessible.

Informed Consent: N/A.

Animal Studies: N/A.

