## Supplementary figures for "The causal relationship between Graves’ disease and type 2 diabetes: Evidence from Bidirectional Mendelian Randomization Study": Supplementary Figures GD ON T2D.docx

**Running Title:** Relationship between Graves' disease and type 2 diabetes

**Authors:**

1. Mahdi Akbarzadeh; Ph.D., Cellular and Molecular Endocrine Research Center, Research Institute for Endocrine Sciences, Shahid Beheshti University of Medical Sciences, Tehran, Iran. Email: .
2. Mahsa Heidari-Foroozan, M.D., Student Research Committee, School of Medicine, Shahid Beheshti University of Medical Sciences, Tehran, Iran. Email: .
3. Samaneh Talebi, Ph.D., Department of Biostatistics, School of Allied Medical Sciences, Shahid Beheshti University of Medical Sciences, Tehran, Iran..
4. Danial Habibi; Ph.D., Department of Biostatistics and Epidemiology, School of Health, and Student Research Committee, School of Health, Isfahan University of Medical Sciences, Isfahan, Iran. Email: .
6. Farideh Neshati; MS.c., Cellular and Molecular Endocrine Research Center, Research Institute for Endocrine Sciences, Shahid Beheshti University of Medical Sciences, Tehran, Iran. Email: .
7. Majid Valizadeh, MD, Obesity Research Center, Research Institute for Endocrine Sciences, Shahid Beheshti University of Medical Sciences, Tehran, Iran..
8. Amir Hossein Ghanooni; MD, Department of Endocrinology, School of Medicine, Iran University of Medical Sciences, Tehran, Iran.
9. Hamid Alavi Majd; Ph.D., Department of Biostatistics, School of Allied Medical Sciences, Shahid Beheshti University of Medical Sciences, Tehran, Iran. Email: .
10. Parisa Riahi; MSc, Cellular and Molecular Endocrine Research Center, Research Institute for Endocrine Sciences, Shahid Beheshti University of Medical Sciences, Tehran, Iran.
11. Fereidoun Azizi; MD, Endocrine Research Center, Research Institute for Endocrine Sciences, Shahid Beheshti University of Medical Sciences, Tehran, Iran. Email: .
12. Mehdi Hedayati; Ph.D., Cellular and Molecular Endocrine Research Center, Research Institute for Endocrine Sciences, Shahid Beheshti University of Medical Sciences, Tehran, Iran. Email: .
13. Maryam Sadat Daneshpour*; Ph.D., Cellular and Molecular Endocrine Research Center, Research Institute for Endocrine Sciences, Shahid Beheshti University of Medical Sciences, Tehran, Iran. Email: .

**Corresponding author:**

**Maryam Sadat Daneshpour** (Ph.D.), Associate Professor. Cellular and Molecular Endocrine Research Center, Research Institute for Endocrine Sciences, Shahid Beheshti University of Medical Sciences. Email: ; P.O. Box: 19395- 4763, 1985717413, Tel: +98 (21) 22432500, Fax: +98 (21) 22402463.

**Table of content:**

Fig. S1. A: Mr_scatter_plot, B: Funnel plot of causal association between Graves' disease and Type 2 diabetes, C: Leave-one-out plot to assess if a single variant is driving the association between Graves' disease and Type 2 diabetes, and D: Forest plot of variant specific inverse variance estimates for causal association between Graves' disease and Type 2 diabetes.

Fig. S2. A: Mr_scatter_plot, B: Funnel plot of causal association between Type 2 diabetes and Graves' disease, C: Leave-one-out plot to assess if a single variant is driving the association between Type 2 diabetes and Graves' disease, and D: Forest plot of variant specific inverse variance estimates for causal association between Type 2 diabetes and Graves' disease.

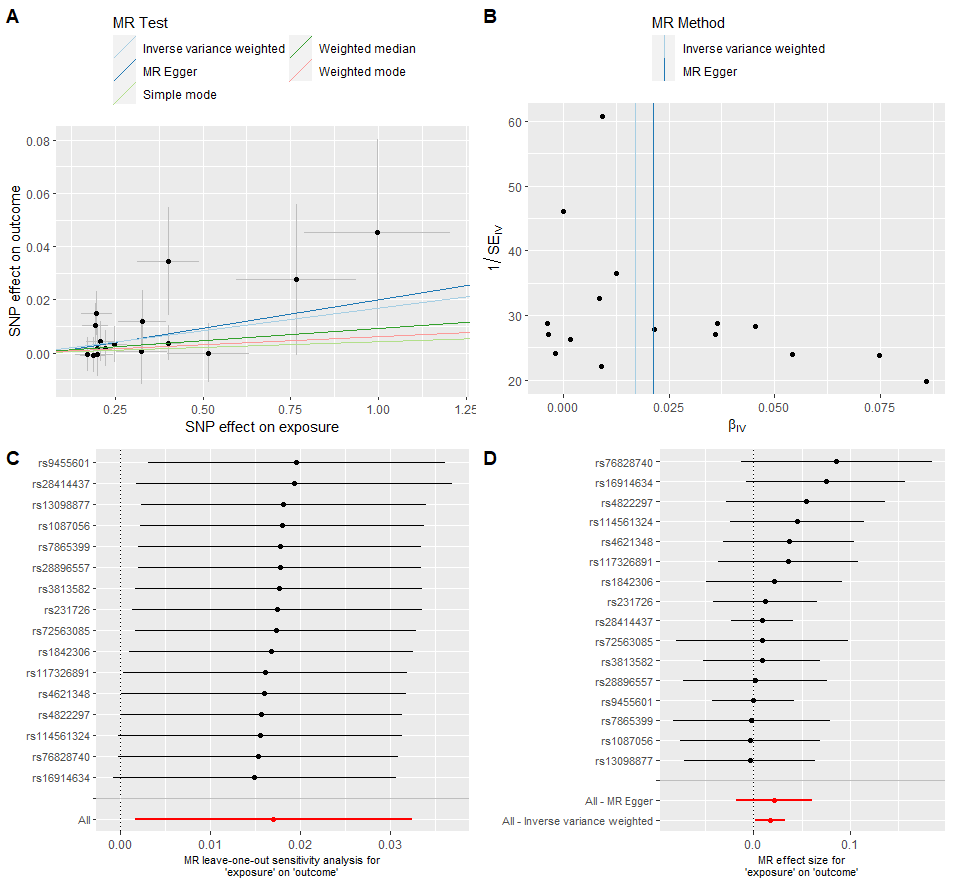

**Fig.** S1. A: Mr_scatter_plot, B: Funnel plot of causal association between Graves' disease and Type 2 diabetes, C: Leave-one-out plot to assess if a single variant is driving the association between Graves' disease and Type 2 diabetes, and D: Forest plot of variant specific inverse variance estimates for causal association between Graves' disease and Type 2 diabetes.

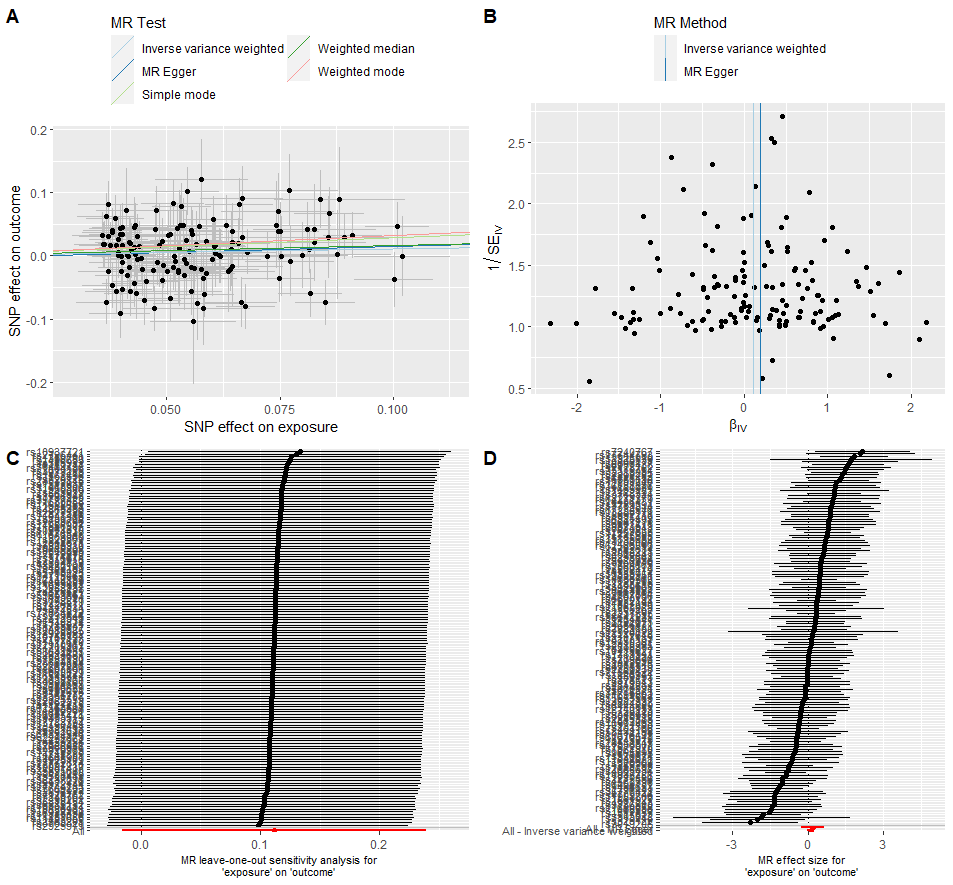

**Fig**. S2. A: Mr_scatter_plot, B: Funnel plot of causal association between Type 2 diabetes and Graves' disease, C: Leave-one-out plot to assess if a single variant is driving the association between Type 2 diabetes and Graves' disease, and D: Forest plot of variant specific inverse variance estimates for causal association between Type 2 diabetes and Graves' disease.
